# Effects of Adolescent Parenting Styles on Adult Cardiovascular Conditions: A Population-Based Cohort Study

**DOI:** 10.64898/2026.08.22.26361103

**Authors:** Nazihah Noor, Josephine Jackisch, Stéphanie Baggio, Stéphane Cullati, Cristian Carmeli

## Abstract

**Purpose:** Family-based interventions are proposed for primordial cardiovascular disease (CVD) prevention, yet which family-environment components to target remains unclear. We quantified effects of parenting styles in adolescence on adult cardiovascular conditions, including hypertension, and whether effects differ by family financial hardship.

**Methods:** Data were from the US National Longitudinal Study of Adolescent to Adult Health (n=4,050). Parenting styles were derived via latent class analysis of adolescent-reported parental responsiveness and demandingness (ages 12–19, 1994–1995). Family financial hardship was based on parent-reported ability to pay bills. CVD and hypertension were assessed via biomarkers and self-report (ages 33–43, 2016–2018). Confounding factors were selected based on a directed acyclic graph; risk differences were estimated using doubly robust inverse-probability-weighted models.

**Results:** Three parenting styles emerged: authoritative (11.1%), permissive (77.9%), and indifferent (11.0%). After 21 years, 33.0% had CVD or hypertension. Whole-population risk differences for permissive and indifferent versus authoritative parenting were -1.0% (95% CI: -5.3, 3.3%) and -1.8% (95% CI: -7.8, 4.2%), respectively. Among families reporting financial hardship, permissive parenting had lower risk (-15.9%, 95%CI: -28.4%, -3.3%), though inconsistent across sensitivity analyses.

**Conclusions:** Adolescent parenting styles had small estimated long-term cardiovascular effects, with no robust evidence of differences by financial hardship.

## 1. Introduction

Cardiovascular diseases (CVDs) and hypertension are leading causes of premature morbidity an^i^d mortality worldwide, with socioeconomically disadvantaged populations bearing a disproportionate burden ^1^. Hypertension is itself a major cardiovascular condition, and a key driver of the atherosclerotic and structural cardiac changes underlying CVDs ^2^. While research has focused on health behaviors as drivers of cardiovascular conditions ^3,4^, growing evidence suggests that family environments during early life shape health trajectories and contribute to the social gradient in cardiovascular outcomes ^5–9^. Accordingly, family-based approaches have been proposed as part of primordial CVD prevention strategies ^9^.

Parenting style is a key, potentially modifiable aspect of the family environment. Parenting styles can shape children’s development ^10^ and have long-term consequences for self-rated health, cognitive function, mental health and mortality in adulthood ^10–12^. Baumrind’s typology conceptualizes parenting along two dimensions: responsiveness (warmth, support) and demandingness (rules-setting, expectations, monitoring), yielding four styles: authoritative (high responsiveness and high demandingness,) authoritarian (low responsiveness and high demandingness), permissive high responsiveness and low demandingness), and indifferent (low responsiveness and low demandingness) ^13,14^. Authoritative parenting is generally considered most beneficial for child development, balancing emotional support with autonomy, and translating into positive psychosocial developmental outcomes and academic achievement ^15–17^.

Despite extensive research on parenting and child development, evidence on long-term effects of parenting styles on cardiovascular outcomes remains limited. Existing research provides several reasons to consider parenting styles during adolescence as potentially relevant for adult cardiovascular conditions. First, specific parenting components (warmth, support, consistent monitoring) have been associated with blood pressure and cardiovascular risk in adolescence and young adulthood ^8,18,19^. Second, several longitudinal studies show parenting styles are associated with causal CVD risk factors including body mass index trajectories, health behaviors and psychosocial wellbeing from adolescence to adulthood ^11,15,16,20–22^. Overall, these findings underscore the need for studies quantifying the long-term effects of parenting styles on CVDs and hypertension.

Additionally, financial hardship may modify these effects by disrupting family functioning and amplifying negative consequences of suboptimal parenting. The Family Stress Model and the Family Investment Model both highlight distinct but interrelated mechanisms by which financial hardship affects parenting and long-term health outcomes. The Family Stress Model emphasizes the psychological and relational strain induced by economic pressures, whereas the Family Investment Model highlights the material and educational investments that are enabled by financial resources ^23,24^. Thus, financial hardship may lead to differential susceptibility, whereby social inequalities in health may arise when disadvantaged individuals experience greater harm from adverse exposures ^25^. Understanding whether family financial hardship modifies long-term effects of parenting styles on cardiovascular outcomes is essential for etiological research and public health practice. From a prevention perspective, evidence of differential susceptibility could inform whether population-wide or targeted family-based interventions are needed for reducing socioeconomic inequality in cardiovascular conditions.

The present study quantifies effects of parenting styles during adolescence on adult CVD and hypertension risk and differential effects by family financial hardship.

## 2. Methods

The study design, variable definitions, and analytic approach were pre-registered in Open Science Framework before data analysis began ^26^. Ethical approval was obtained from the Ethics Commission for Research on Human Beings of the Vaud Canton (CER-VD, file No. 2026-01018).

### 2.1. Study and analytic populations

Our target population is adolescents living with parents in the United States. We used data from the National Longitudinal Study of Adolescents to Adult Health (Add Health), a nationally representative longitudinal study that followed adolescents from early adolescence into adulthood ^27^. Baseline data were collected through in-home interviews with both adolescents and their parents in 1994 and 1995 (Wave I; ages 12–21).

Figure 1 displays the participant flowchart. The study population included 19,189 individuals aged 12–19. For the descriptive analysis, we further excluded respondents missing parenting items (n = 812) or Wave I design information (n = 1,556), yielding 16,821 participants. The causal analytic sample comprised 4,050 individuals who participated in the Wave V (2016– 2018; ages 33–43) biomarker exam, had complete sampling design information, and complete parent-reported family financial hardship during adolescence. Survey weights accounting for unequal selection probabilities to biomarker collection at Wave V and attrition from Wave I to V were applied to obtain nationally representative estimates. Individuals with missing parenting items or family financial hardship were excluded because we hypothesized missingness not at random for these variables.

**Figure 1.**
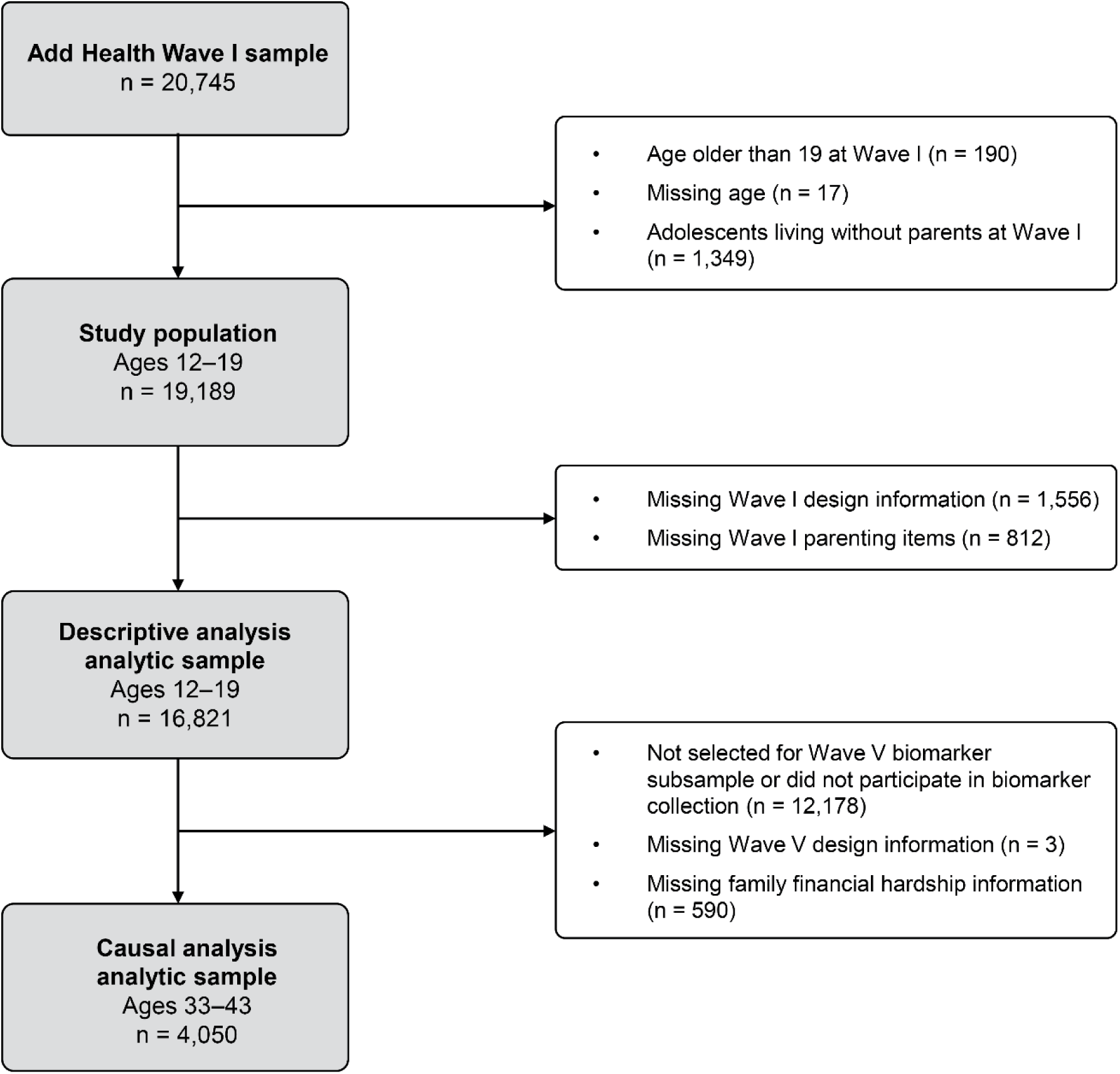
Flowchart of participant selection.

### 2.2. Measures

#### 2.2.1. Exposure: Parenting style

Parenting style during adolescence was derived via latent class analysis (see Section 2.3) of adolescent-reported parental responsiveness and demandingness at Wave I, with variables described in Supplementary Material A.

Responsiveness was assessed through five pre-specified items capturing perceived parental warmth and involvement. These included how close adolescents felt to their parents, how much they believed their parents cared about them, and their perceptions of warmth and affection, communication quality, and overall relationship satisfactions. Responses were given on five-point scales ranging from 1 (“not at all”) to 5 (“very much”) for closeness and caring, and from 1 (“strongly agree”) to 5 (“strongly disagree”) for warmth, communication, and relationship satisfaction.

Demandingness was assessed through seven pre-specified items reflecting parenting control over key aspects of the adolescent’s daily lives, including curfew, friendships, clothing, hours of television watched, which television programs were watched, bedtime hours and food choices. Each item asked whether the adolescent was allowed to make their own decisions (yes/no).

#### 2.2.2. Outcome: CVDs and hypertension

The outcome was defined as the presence of any of the following conditions in adulthood, measured at Wave V: stroke, heart failure, heart disease, and atrial fibrillation (collectively referred to as CVDs), as well as hypertension. Participants were considered to have the outcome if they reported any of these conditions. A separate analysis was also conducted with hypertension as the sole outcome.

Stroke, heart failure, atrial fibrillation, and heart disease were identified through self-reported previous diagnosis by a healthcare professional, with heart disease additionally including a self-reported history of heart surgery for clogged coronary arteries. Hypertension was identified through blood pressure readings (systolic blood pressure ≥140 mmHg or diastolic blood pressure ≥90 mmHg), self-reported diagnosis of hypertension by a healthcare professional, or self-reported use of antihypertensive medication (Supplementary Material B).

#### 2.2.3. Effect modifier: Family financial hardship

Family financial hardship was assessed from the parent questionnaire at Wave I using the question, “Do you have enough money to pay your bills?” (yes/no). Adolescents whose parents responded “no” were classified as having family financial hardship.

#### 2.2.4. Confounding factors

Confounding factors were selected using a directed acyclic graph, informed by existing literature and the authors’ expert knowledge, which was specified a priori in the study protocol^26^ and is presented in Supplementary Material C. The minimally sufficient adjustment set included age at baseline, sex, race/ethnicity, history of chronic disease, neighborhood deprivation, family structure, family-level health behaviors, parental age at birth, parental race/ethnicity, parental psychosocial stress, parental occupation, parental education, and migration background. All confounding factors were measured at Wave I. Detailed definitions and operationalization of all confounders are provided in Supplementary Material D.

### 2.3. Statistical analysis

We conducted descriptive and causal analyses. The descriptive analysis identified parenting styles based on parental responsiveness and demandingness, which informed the causal analysis quantifying their effects on the outcome.

#### Descriptive analysis

We first conducted a confirmatory factor analysis to assess the dimensional structure and internal consistency of the parenting responsiveness and demandingness items. McDonald’s omega was used to evaluate scale reliability ^28^.

We then conducted a latent class analysis (LCA) to empirically identify patterns of parenting styles. Models specifying one to five latent classes were estimated using weighted ordinal logistic models in Stata 18. The optimal number of classes was determined based on standard model fit criteria (Supplementary Material J), while also considering class interpretability. Participants were assigned to their most likely class based on posterior probabilities, and these classifications were used as exposure in the causal analysis.

#### Causal analysis

We estimated the average causal effects (ACEs) of the identified parenting styles on adult CVD and hypertension, as well as the differences in these effects by family financial hardship (conditional ACEs). Each ACE represents the marginal difference in potential outcomes between two counterfactual scenarios: one in which adolescents are exposed to a sub-optimal parenting style, and one in which they are exposed to an optimal parenting style. Conditional ACEs were estimated separately for each hardship stratum. Estimated associations were measured via risk differences because of their direct interpretability in relation to disease prevalence and potential intervention impacts ^29^. A doubly robust approach with inverse-probability weighting with outcome regression was implemented (Supplementary Material F). Stabilized inverse probability weights were computed from an exposure regression model and truncated at the 2.5^th^ and 97.5^th^ percentiles to limit the influence of extreme values and satisfy the positivity assumption ^30^. Balance of measured confounders after weighting was assessed using standardized mean differences (Supplementary Material E). Interpretation of the estimated adjusted associational risk differences as causal effects relied on assumptions of no residual confounding, consistency, and positivity.

Estimates were averaged across 100 multiple imputed datasets and compatibility intervals (CIs) were derived from 1,000 bootstrapped sampling designs using the Rao-Wu-Yue-Beaumont method (Supplementary Material G). This analysis was performed in R version 4.5.1.

### 2.4. Sensitivity analyses

First, we assessed the sensitivity of the adjusted risk differences to an alternative operationalization of parenting style, using a predefined composite approach that dichotomizes parental responsiveness and demandingness and combines them into the four Baumrind’s categories ^31^.

Second, we assessed potential residual confounding using a negative control exposure, parental seatbelt use while driving. This exposure was selected as a proxy for hard-to-measure parental characteristics such as self-regulation, risk aversion, and risk perception, which may influence parenting style and are also related to psychosocial stress ^32^. We assumed parental seatbelt use does not directly affect offspring CVD or hypertension risk but may share unmeasured or partially measured common causes with parenting style. Specifically, although we adjusted for parental psychosocial stress using self-reported general happiness, this measure captures only part of the construct. Therefore, an adjusted association between parental seatbelt use and offspring outcomes would suggest residual confounding by such partially measured characteristics rather than a causal effect ^33^.

Third, we used parental education as an alternative effect modifier. While parental education may be more indicative of cultural rather than purely economic capital, it is strongly correlated with parental income/financial strain in the US, and it is more stable across the life course ^34^.

## 3. Results

### 3.1. Causal analysis sample characteristics

Table 1 summarizes the characteristics of adolescents in our causal analytic sample, with weighted percentages provided in parentheses. Most respondents were in mid-to-late adolescence (72.0%); 50.4% were female. The majority were White (70.0%), followed by Black (14.8%), Hispanic (11.0%) and other racial/ethic groups (3.9%). Most parents had high education (67.0%). Most adolescents came from families without financial hardship (83.8%). After an average follow-up of 21.3 years, 33.0% had CVD or hypertension, and 31.8% had hypertension (with or without CVD). Adolescents from families with financial hardship had a higher prevalence of CVD and hypertension compared to advantaged peers (38.8% vs. 30.9%, respectively, Supplementary Material H).

**Table 1.**
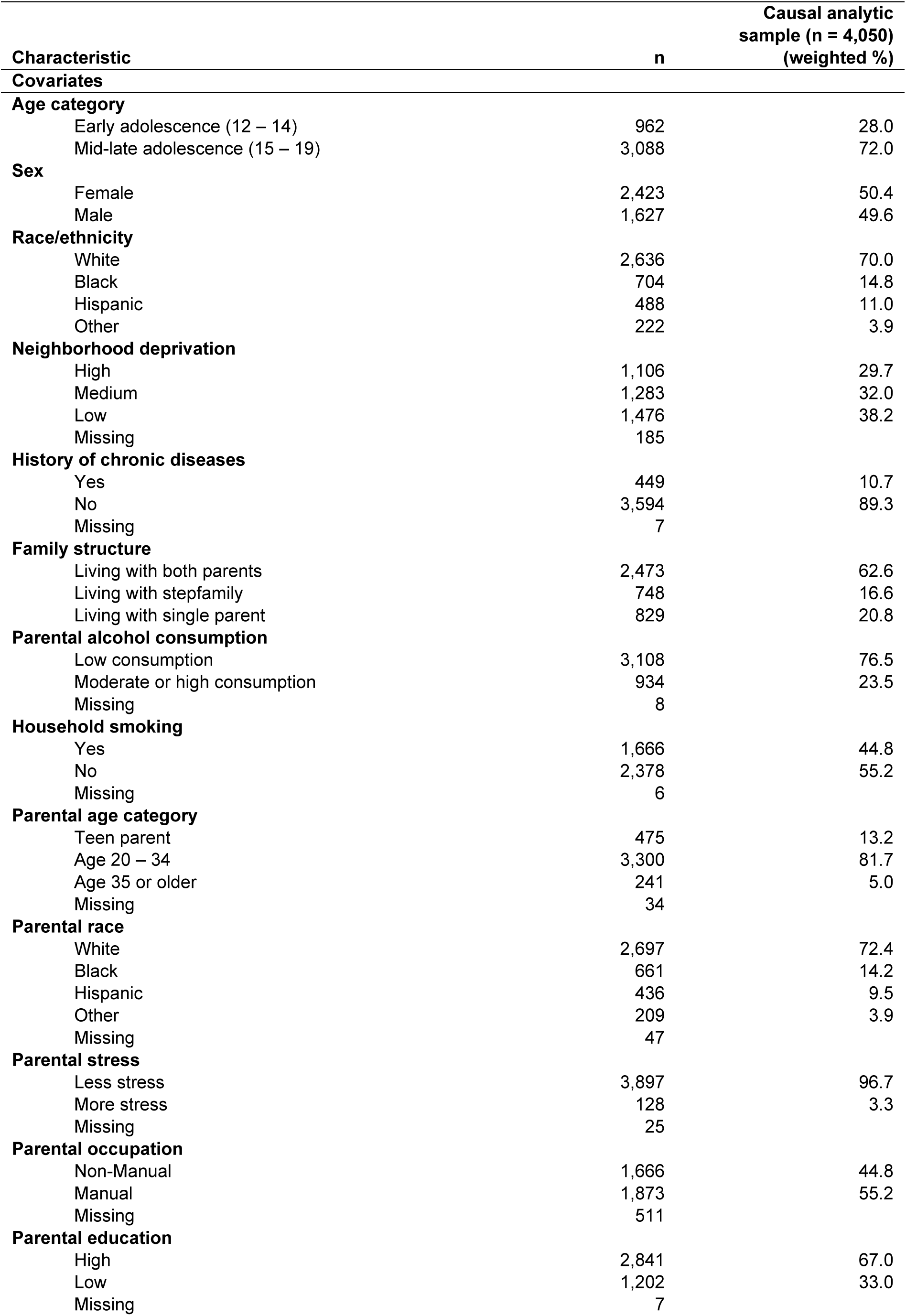
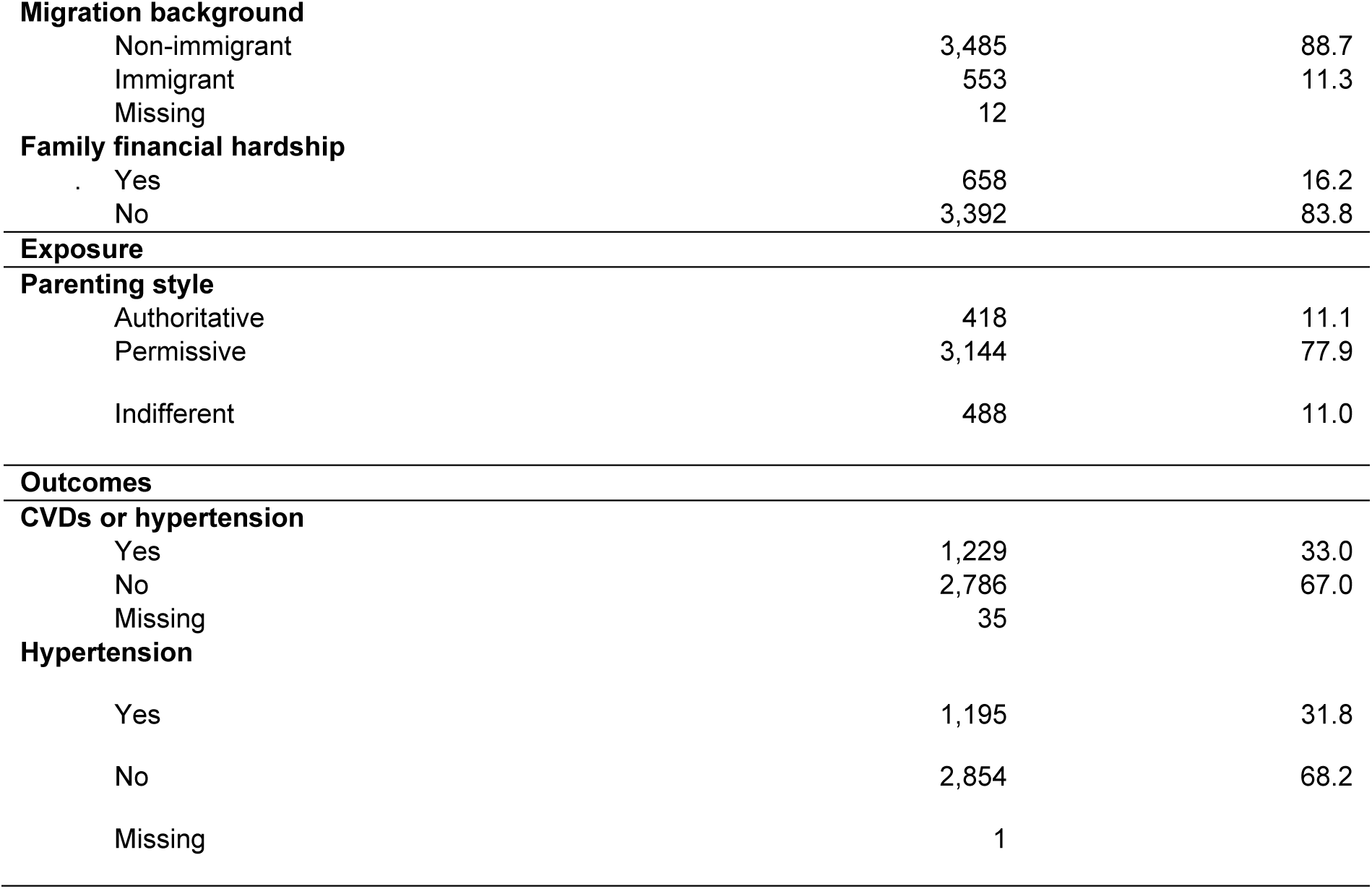
Summary of causal analytic sample characteristics.

| **Characteristic** | **n** | **Causal analytic sample (n = 4,050) (weighted %)** |
| --- | --- | --- |
| **Covariates** |  |  |
| **Age category** |  |  |
| Early adolescence (12 – 14) | 962 | 28.0 |
| Mid-late adolescence (15 – 19) | 3,088 | 72.0 |
| **Sex** |  |  |
| Female | 2,423 | 50.4 |
| Male | 1,627 | 49.6 |
| **Race/ethnicity** |  |  |
| White | 2,636 | 70.0 |
| Black | 704 | 14.8 |
| Hispanic | 488 | 11.0 |
| Other | 222 | 3.9 |
| **Neighborhood deprivation** |  |  |
| High | 1,106 | 29.7 |
| Medium | 1,283 | 32.0 |
| Low | 1,476 | 38.2 |
| Missing | 185 |  |
| **History of chronic diseases** |  |  |
| Yes | 449 | 10.7 |
| No | 3,594 | 89.3 |
| Missing | 7 |  |
| **Family structure** |  |  |
| Living with both parents | 2,473 | 62.6 |
| Living with stepfamily | 748 | 16.6 |
| Living with single parent | 829 | 20.8 |
| **Parental alcohol consumption** |  |  |
| Low consumption | 3,108 | 76.5 |
| Moderate or high consumption | 934 | 23.5 |
| Missing | 8 |  |
| **Household smoking** |  |  |
| Yes | 1,666 | 44.8 |
| No | 2,378 | 55.2 |
| Missing | 6 |  |
| **Parental age category** |  |  |
| Teen parent | 475 | 13.2 |
| Age 20 – 34 | 3,300 | 81.7 |
| Age 35 or older | 241 | 5.0 |
| Missing | 34 |  |
| **Parental race** |  |  |
| White | 2,697 | 72.4 |
| Black | 661 | 14.2 |
| Hispanic | 436 | 9.5 |
| Other | 209 | 3.9 |
| Missing | 47 |  |
| **Parental stress** |  |  |
| Less stress | 3,897 | 96.7 |
| More stress | 128 | 3.3 |
| Missing | 25 |  |
| **Parental occupation** |  |  |
| Non-Manual | 1,666 | 44.8 |
| Manual | 1,873 | 55.2 |
| Missing | 511 |  |
| **Parental education** |  |  |
| High | 2,841 | 67.0 |
| Low | 1,202 | 33.0 |
| Missing | 7 |  |
| **Migration background** |  |  |
| Non-immigrant | 3,485 | 88.7 |
| Immigrant | 553 | 11.3 |
| Missing | 12 |  |
| **Family financial hardship** |  |  |
| Yes | 658 | 16.2 |
| No | 3,392 | 83.8 |
| **Exposure** |  |  |
| **Parenting style** |  |  |
| Authoritative | 418 | 11.1 |
| Permissive | 3,144 | 77.9 |
| Indifferent | 488 | 11.0 |
| **Outcomes** |  |  |
| **CVDs or hypertension** |  |  |
| Yes | 1,229 | 33.0 |
| No | 2,786 | 67.0 |
| Missing | 35 |  |
| **Hypertension** |  |  |
| Yes | 1,195 | 31.8 |
| No | 2,854 | 68.2 |
| Missing | 1 |  |

Baseline characteristics were similar between participants included in the causal analytic sample and those eligible for the Wave V biomarker assessment prior to exclusion for missing financial hardship and parenting style items (Supplementary Material I).

### 3.2. Confirmatory factor analysis

Based on factor loadings and reliability indices (McDonald’s ω = 0.89 for responsiveness; ω = 0.75 for demandingness), six of the seven pre-specified demandingness items were retained, with curfew dropped due to low factor loadings, and all five pre-specified responsiveness items were retained for the latent class analysis (Supplementary Material J).

### 3.3. Latent class analysis of parenting styles

Based on model fit indices and interpretability (Supplementary Material K), a four-class model was selected. As shown in Figure 2, one class was characterized by high responsiveness and high demandingness, which we defined as the authoritative parenting group (11.1% of the weighted sample, Class 2). Two classes (Classes 1 and 4) exhibited high responsiveness combined with comparatively lower demandingness. Although these two classes differed slightly in responsiveness, they did not differ meaningfully in demandingness, and were therefore aggregated into a single permissive parenting category (77.9%). One class showed low responsiveness and low demandingness, characterized as the indifferent parenting group (11.0%, Class 3). No class consistent with an authoritarian style was identified.

**Figure 2.**
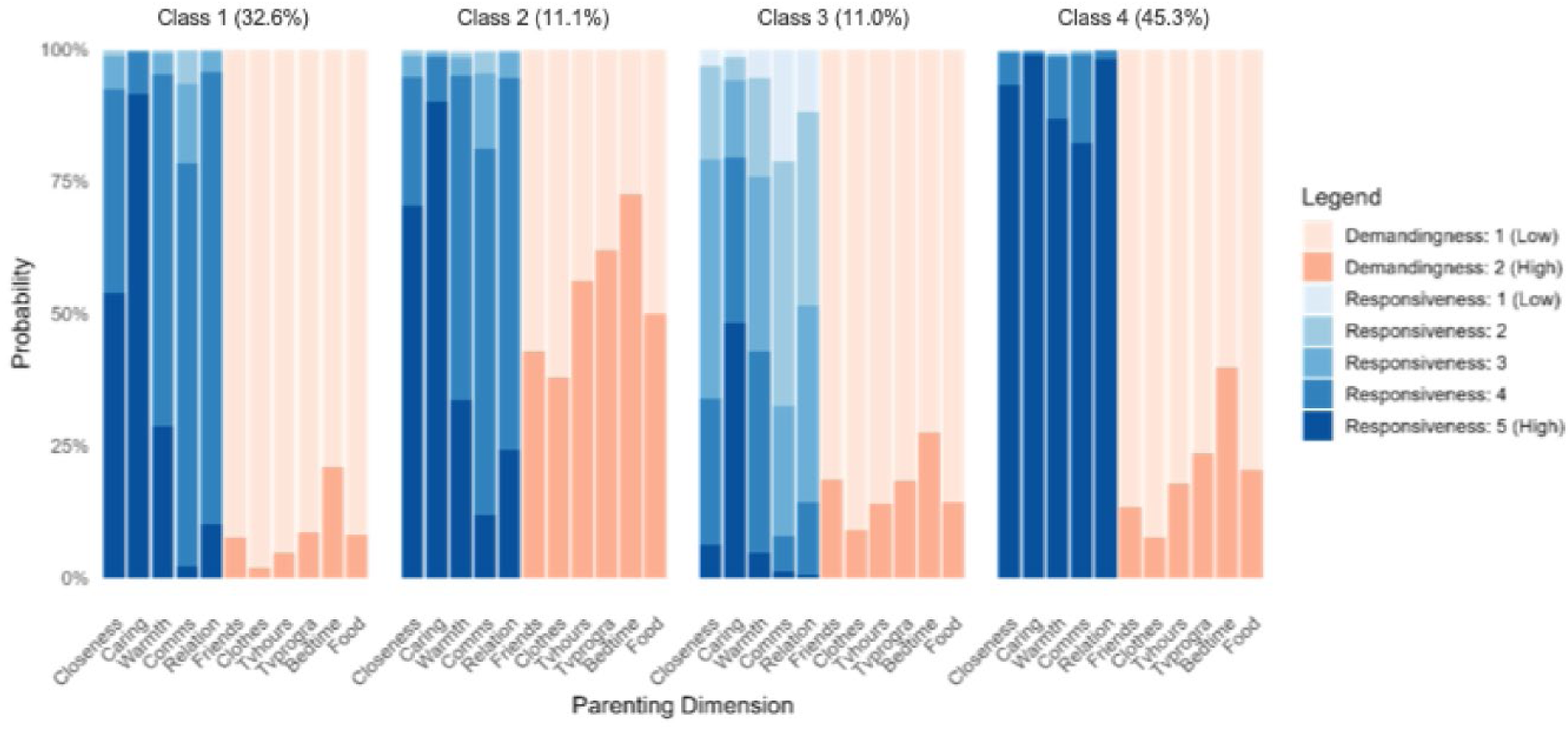
Item-response probabilities by latent class for five responsiveness items (perceived closeness, caring, warmth, communication quality and relationship satisfaction) and six demandingness items (control over friends, clothing, television hours, television programs, bedtime and food). Classes are labeled post-hoc as authoritative (class 2), permissive (classes 1 and 4) and indifferent (class 3).

### 3.4. Average and conditional causal effects of parenting styles on CVD and hypertension

Risk differences for permissive and indifferent parenting compared with authoritative parenting were of small magnitude (below 5 percentage points) in the whole-population sample (Figure 3). When stratified by family financial hardship, a distinct pattern emerged for permissive parenting: risk was lower among families experiencing hardship but not among those without, with an effect modification of -18.2% (95% CI: -32.4%, -3.9%). For indifferent parenting, the effect modification estimate was -7.8% (95% CI: -23.7%, 8.2%), with the wide confidence interval precluding any conclusion about differential associations. Risk differences with hypertension as the sole outcome aligned with those for the combined outcome (Supplementary Material L).

**Figure 3.**
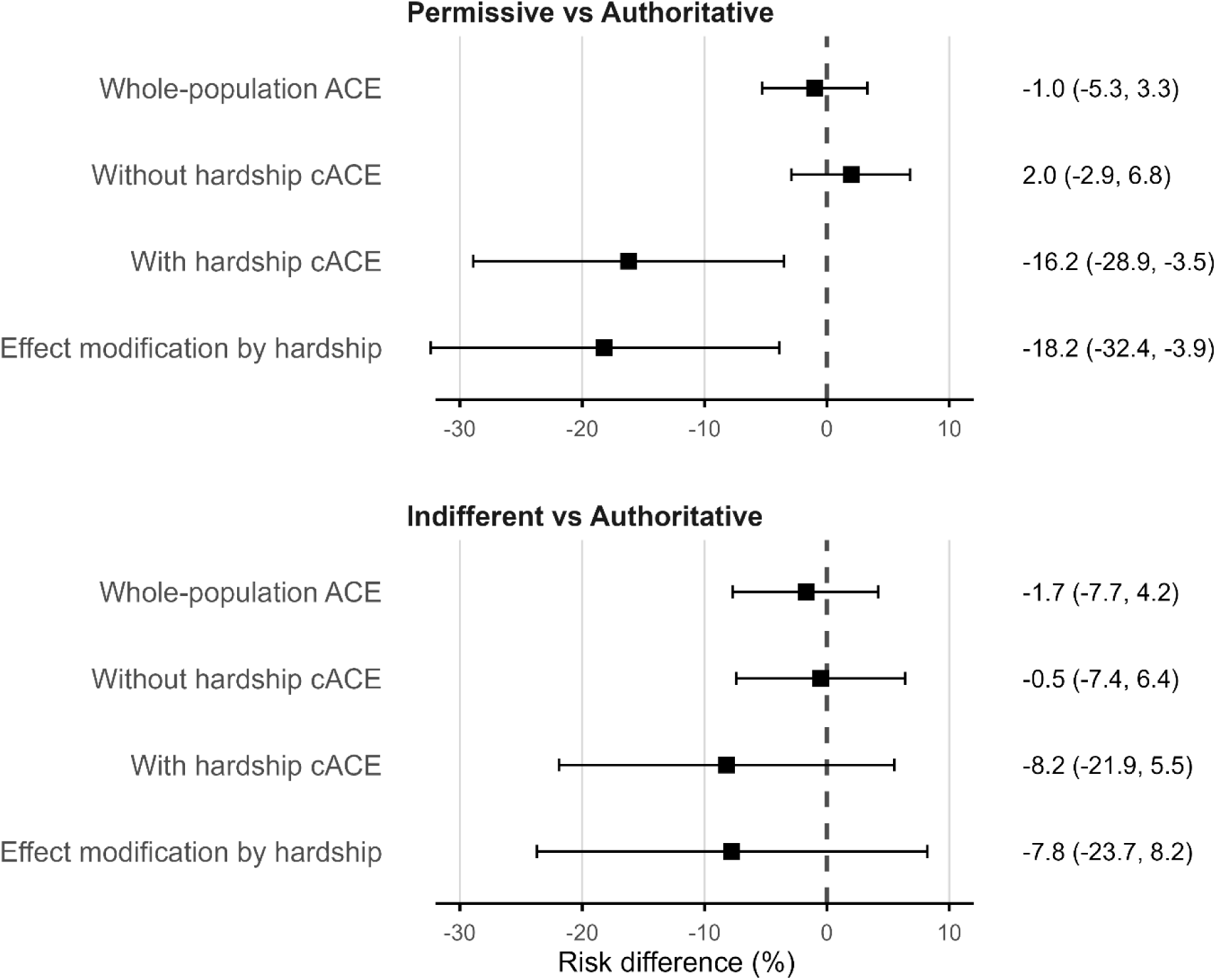
Estimated whole-population average causal effect (ACE), conditional average causal effects (cACE, among adolescents without and with family financial hardship), and effect modification by hardship (difference between the two cACEs), for permissive vs. authoritative (top panel) and indifferent vs. authoritative parenting (bottom panel). Squares represent adjusted associational risk differences (%); their interpretation as causal effects relies on the assumptions stated in Methods. Error bars indicate 95% compatibility intervals. Point estimates and 95% compatibility intervals are also reported in the right-hand column.

### 3.5. Sensitivity analyses

When parenting styles were defined using median splits of responsiveness and demandingness, risk differences aligned with those observed when using LCA-based parenting styles (Supplementary Material M).

The negative control analysis showed whole-population and stratum-specific risk differences of similar magnitude (Supplementary Material M), suggesting the estimated effect modification for permissive parenting may reflect residual confounding rather than causation^35^.

When considering parental education as the effect modifier, estimated effect modifications were in the opposite direction compared to family financial hardship (Supplementary Material M).

## 4. Discussion

In this 21-year follow-up of a nationally representative US cohort, adjusted associational risk differences between suboptimal parenting styles and adult CVD or hypertension were mostly small in magnitude. The association between permissive parenting and lower CVD/hypertension risk among families experiencing financial hardship was inconsistent across sensitivity analyses and may reflect residual confounding rather than a genuine protective effect. For indifferent parenting, there was no clear evidence that associations differed by financial hardship.

Previous research has examined related constructs such as parental relationships, warmth, support and supervision during adolescence, but few studies have quantified how distinct parenting styles relate to adult CVDs and hypertension. Our small whole-population risk differences are broadly consistent with the modest magnitude of previous findings. Doom et al. (2016), using Add Health data, reported that higher maternal support in adolescence was associated with a 0.5% lower predicted 30-year CVD risk scores at ages 24–34 years among over 11,000 participants ^18^. Differences in the exposure definition (maternal warmth vs. parenting style typologies), outcome measurement (predicted risk vs. actual CVD/hypertension), and analytic approach (covariate adjustment vs our doubly robust approach) may explain variation in findings.

The absence of an authoritarian class in our LCA is also noteworthy. Two prior Add Health studies identified four distinct parenting styles aligned with Baumrind’s typology ^21,31^, using different methods: Driscoll et al. (2007) employed median splits on warmth and control, while Fuemmeler et al. (2012) used factor mixture modeling. In our sensitivity analysis replicating Driscoll et al.’s approach, risk differences remained consistent with those obtained in our main analysis, indicating our associational estimates are not sensitive to the parenting style identification method.

Several factors may explain the small effect sizes. First, parenting style typologies may obscure effects of specific traits (e.g. warmth, support or supervision individually). Second, some longitudinal studies suggest adult socioeconomic conditions and health behaviors can mitigate the impact of parenting on cardiovascular outcomes ^36,37^. Thus, the long-term effects of parenting styles during adolescence may be attenuated by exposures accumulated across adulthood, partly explaining our small effect sizes.

Beyond whole-population effects, we examined differential effects by family financial hardship. In the main analysis, permissive parenting was associated with lower CVD or hypertension risk among families experiencing financial hardship but not among those without, suggesting differential susceptibility ^25^. However, this differential association was inconsistent across sensitivity analyses and could reflect residual confounding. For indifferent parenting, the confidence interval for effect modification was wide, precluding any conclusion about differential susceptibility. Overall, these findings provide limited evidence of differential susceptibility to parenting styles by family financial hardship, and should be interpreted cautiously.

Our study has limitations. Despite comprehensive confounding adjustment guided by a causal model, unmeasured or residual confounding cannot be entirely excluded, as suggested by the negative control analysis. Our measurement of demandingness relied on binary response options, which may have captured this dimension imperfectly by restricting variability. Additionally, the absence of sizeable effects could be due to insufficient measurement sensitivity. For example, the Parenting Styles and Dimensions Questionnaire provides multi-item scales validated to capture Baumrind’s typologies ^38^, but such instruments are often infeasible in large population-based surveys. The consistency assumption may be violated given how the exposure is defined. Each latent class aggregates heterogeneous parenting configurations, so our estimates average over an unspecified distribution of versions of the exposure. Moreover, parenting was reported by adolescents, capturing perceived rather than observed parental behavior; whether the relevant causal agent is parental behavior or the adolescent’s experience of it remains unresolved. Our findings therefore capture long-term consequences of broadly characterized parenting environments rather than specific parenting interventions, which warrant separate evaluation. Finally, model miss-specification could have biased our estimates, however we mitigated this possibility by implementing a doubly robust estimator.

Our study has notable strengths. It leverages a large, nationally representative longitudinal dataset with over 20 years of follow up. The combination of LCA and causal inference methods provides a rigorous framework for estimating causal effects under explicitly stated identifying assumptions.

In this nationally representative cohort, parenting styles during adolescence showed small estimated effects on cardiovascular conditions at ages 33–43, with no robust evidence of differences by family financial hardship. Future research using more granular parenting measures, or exposures defined closer to actual interventions, would help clarify whether early family environments meaningfully shape long-term cardiovascular risk.

## Data Availability

The Add Health data are available in public‑use and restricted‑use formats. Restricted‑use data, including the full cohort, require an approved contract. Information on accessing Add Health data is available at https://addhealth.cpc.unc.edu/data/

## Acknowledgements

We thank Kathleen Mullan Harris for feedback on this study protocol. This research uses data from Add Health, a program project directed by Kathleen Mullan Harris and designed by J. Richard Udry, Peter S. Bearman, and Kathleen Mullan Harris at the University of North Carolina at Chapel Hill, and funded by grant P01-HD31921 (Harris) from the Eunice Kennedy Shriver National Institute of Child Health and Human Development, with cooperative funding from 23 other federal agencies and foundations. No direct support was received from grant P01-HD31921 for this analysis.

