## Supplementary Material A for "Effects of Adolescent Parenting Styles on Adult Cardiovascular Conditions: A Population-Based Cohort Study"

### A. Add Health Questions Used for Parental Style

Parental responsiveness and demandingness were assessed using adolescent-reported items at Wave I. The exact questionnaire items are listed below.

**Supplementary Table 1. Add Health survey variables used to identify parental style items.**

| Parenting dimension | Parenting item | Variable name | Survey question |
|---|---|---|---|
| Responsiveness | Closeness | H1WP9 | How close do you feel to your mother? (1 = not at all, 5 = very much) |
| | Caring | H1WP10 | How much do you think she cares about you? (1 = not at all, 5 = very much) |
| | Warmth | H1PF1 | Your mother is warm and loving toward you. (1 = strongly agree, 5 = strongly disagree) |
| | Communication quality | H1PF4 | You are satisfied with the way you and your mother communicate. (1 = strongly agree, 5 = strongly disagree) |
| | Relationship satisfaction | H1PF5 | You are satisfied with your relationship with your mother. (1 = strongly agree, 5 = strongly disagree) |
| Demandingness | Curfew | H1WP1 | Are you allowed to decide what time you must be home on weekend nights? (0 = no, 1 = yes) |
| | Friends | H1WP2 | Are you allowed to decide who you hang out with? (0 = no, 1 = yes) |
| | Clothes | H1WP3 | Are you allowed to decide what clothes you wear? (0 = no, 1 = yes) |
| | Television hours | H1WP4 | Are you allowed to decide how many hours of television you watch? (0 = no, 1 = yes) |
| | Television programs | H1WP5 | Are you allowed to decide which television programs you watch? (0 = no, 1 = yes) |
| | Bedtime | H1WP6 | Are you allowed to decide what time you go to bed on weeknights? (0 = no, 1 = yes) |
| | Food | H1WP7 | Are you allowed to decide what foods you eat? (0 = no, 1 = yes) |

#### B. Hypertension classification

Hypertension status was based on an Add Health constructed variable for joint classification of hypertension, which flags respondents meeting at least one of the following criteria:

1. blood pressure classified as stage 1 or 2 hypertension using thresholds from the Seventh Report of the Joint National Committee on Prevention Detection, Evaluation, and Treatment of High Blood Pressure;
2. self-reported prior diagnosis of high blood pressure; or
3. reported use of antihypertensive medication in the past four weeks.

Blood pressure was measured three times by trained field examiners using a Microlife BP3MC1-PC-IB oscillometric monitor, and the average of the second and third readings was used to derive systolic and diastolic measures [1].

#### C. Directed Acyclic Graph

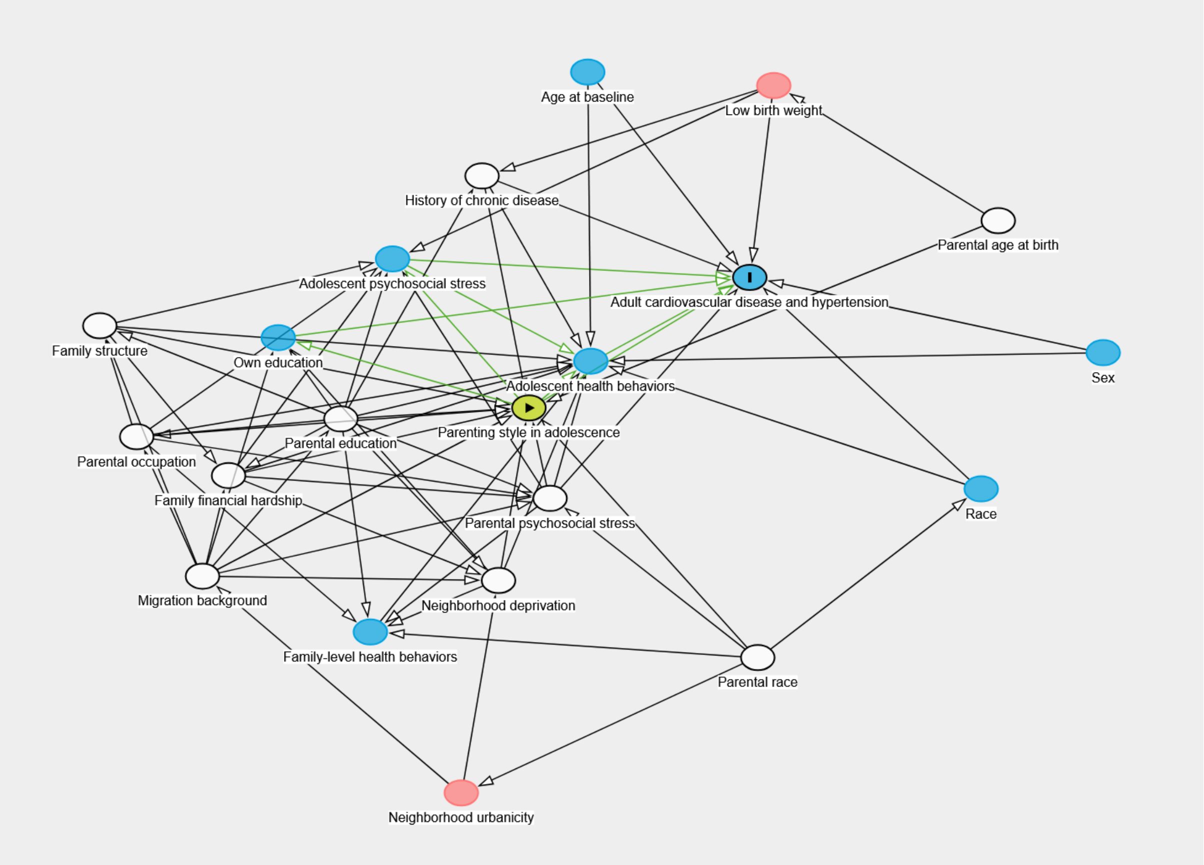

**Supplementary Figure 1.** Causal analytic model portrayed as a directed acyclic graph. The exposure is marked with a triangle in a green circle and the outcome with an I in a blue circle. Variables in white nodes mark the minimal sufficient adjustment set. The graph was drawn with DAGitty [2] and can be found here: https://dagitty.net/mDEYSZoEs

#### D. Operationalization of Confounding Factors

Age at baseline survey was measured at Wave I via self-reported date of birth and categorized into early adolescence (12 – 14 years) and mid-late adolescence (15 – 19 years).

Sex at birth was self-reported at Wave I and categorized as male or female.

Race/ethnicity was measured through self-report and interviewer-identified at Wave I and categorized into (non-Hispanic) White, (non-Hispanic) Black, Hispanic and (non-Hispanic) Other.

History of chronic disease was derived from self-reported physician diagnoses of hypertension, diabetes, stroke, heart failure, cancer or asthma occurring before the baseline interview.

Neighborhood deprivation was measured using an Add Health constructed neighborhood disadvantage score linked to respondents' residential addresses at baseline, based on indicators including proportion of female-headed households, individuals living below the poverty threshold, individuals receiving public assistance, adults with less than a high school education, and adults who were unemployed. The score was standardized to have a mean of zero and standard deviation of one, and then categorized into tertiles (high, medium or low deprivation) (Belsky, Domingue et al. 2020).

Family structure during adolescence was based on adolescents' self-reported household composition at Wave I, which recorded the relationship of each household member to the respondent. Add Health constructed detailed family structure indicators categorizing whether respondents lived with both biological (or adoptive) parents, a stepparent, or a single parent (Harris 1999). For the present analysis, family structure was classified as living with both parents (biological or adoptive), living in a step-family, or living with a single parent.

Family-level health behaviors included parental alcohol consumption and household smoking. Parental alcohol consumption was reported by the responding parent and dichotomized as low (never or less than once per month) versus moderate/high (more than 2 – 3 times per month). Household smoking was coded as yes if any household member smoked.

Parental age at birth was derived from subtracting the adolescent's age at baseline from the responding parent's age, with implausible values (parents younger than 12 or mothers older than 50 years) coded as invalid. Parental age at birth was categorized into teenage parents (< 20 years), 20 – 34 years, and 35 years or older.

Parental race/ethnicity was determined from the responding parent’s self-report and categorized as (non-Hispanic) White, (non-Hispanic) Black, Hispanic and (non-Hispanic) Other.

Parental psychosocial stress was proxied using the parent’s self-reported general happiness (“in general, are you happy?” yes/no). Parents responding “no” were categorized as having more psychosocial stress.

Parental occupation was based on adolescent-reported information on the current occupation of resident parents. Occupations were classified as manual (service, craft, agricultural or construction work) or non-manual (professional, managerial, technical or office work).

Parental education was coded as “high” if at least one parent had graduated from a college or university, otherwise “low”, using information from both the parent survey and the adolescent survey in Wave I.

Migration background was dichotomized as immigrant (first or second generation) or non-immigrant. First-generation immigrations were respondents born outside the US, while second-generation immigrants were those born in the US with at least one foreign-born parent.

#### E. Standardized Mean Differences

We assessed covariate balance across parenting style groups by comparing standardized mean differences (SMDs) before and after applying inverse probability weighting. The SMDs were calculated for each of the 100 imputed datasets generated using multiple imputation by chained equations (mice package in R version 4.5.1) and then averaged across imputations. The SMDs were estimated using the cobalt package in R version 4.5.1, following weighting with stabilized inverse probability weights. The weighting model adjusted for measured confounders including baseline age, sex, race/ethnicity, neighborhood deprivation, family structure, parental education, parental occupation, parental alcohol use, household smoking, parental age at birth, parental race/ethnicity, parental stress, migration background and family financial hardship. Consistent with recommendations for multi-category treatments, we considered SMD values below 0.1 to indicate adequate balance. As shown in Supplementary Figure 2 below, after weighting, SMDs were all below 0.1, which is acceptable for multinomial propensity score models [3].

**Supplementary Figure 2.** Love plots of the covariance balance before and after inverse-probability of treatment weighting by treatment variables across the 100 imputed samples. Dashed line corresponds to abs(SMD) = 0.1, values below are usually indicative of an achieved good balance.

#### F. Inverse Probability Weights

We used inverse probability of the exposure weights in the causal analysis. Weights were calculated from a multinomial model relating the three-category parenting style exposure (dependent variable) to measured confounding factors (independent variables), specified as additive and linear without product terms. Final weights were stabilized and truncated at the $2.5^{th}$ and $97.5^{th}$ percentiles to minimize bias arising from few participants having extreme weights. To assess the weight distribution, we checked whether the means were roughly 1 and that trimming removed excessively large values. We also assessed whether the weighted population corresponded to a population in which measured confounding factors were equally distributed across the exposure groups. This was done by comparing standardized mean differences before and after weighing (see previous section).

We specified a marginal structural outcome model for each outcome as a function of parenting style, family financial hardship, their product term and the measured confounding factors included in the exposure model. As shown by Robertson et al., this guarantees our effect or differential effect estimates are double-robust [4].

#### G. Imputation Models

To deal with missingness in variables that we hypothesized were missing at random, we implemented multiple imputations by chained equations using the mice package in R [5]. For each bootstrap sample, we computed 100 imputed datasets and then pooled the estimates by averaging. Predictors for the imputation included all complete covariates and several auxiliary variables that capture additional aspects of the adolescent's family and social environment: availability of cigarettes in the household reported by the adolescent or interviewer (variables H1TO50 and H1IR23 in Add Health), perceived neighborhood safety (H1NB5), perceived social acceptance (H1PF35), unmet medical care due to cost (H1GH26 and H1GH27I) and parental disability (H1RM10, H1RF10 and PA18).

#### H. Inequalities in Exposures and Outcomes

**Supplementary Table 2. Distribution of exposures and outcomes by family financial hardship**. Percentages (%) are weighted.

| | Family financial hardship | | | |
|---|---|---|---|---|
| | Yes | | No | |
| | n | % | n | % |
| **Parenting style** | | | | |
| Authoritative | 76 | 11.6 | 342 | 10.1 |
| Permissive | 477 | 72.5 | 2667 | 78.6 |
| Indifferent | 105 | 16.0 | 383 | 11.3 |
| **CVDs or hypertension** | | | | |
| Yes | 238 | 38.8 | 991 | 30.9 |
| No | 414 | 61.2 | 2372 | 69.1 |
| Missing | 6 | | 29 | |

### I. Comparison of Analytic and Eligible Samples

**Supplementary Table 3. Baseline characteristics of the eligible Wave V biomarker sample and final causal analytic sample.** Percentages (%) are weighted.

| Characteristic | Eligible W5 biomarker sample (n = 4,640) | Final causal analytic sample (n = 4,050) |
|---|---|---|
| **Age category** | | |
| Early adolescence (12 – 14) | 27.1% | 28.0% |
| Mid–late adolescence (15 – 19) | 72.9% | 72.0% |
| **Sex** | | |
| Female | 50.8% | 50.4% |
| Male | 49.2% | 49.6% |
| **Race/ethnicity** | | |
| White | 68.4% | 70.0% |
| Black | 15.7% | 14.8% |
| Hispanic | 11.6% | 11.0% |
| Other | 4.3% | 3.9% |
| **Neighborhood deprivation** | | |
| High | 30.5% | 29.7% |
| Medium | 32.3% | 32.0% |
| Low | 37.1% | 38.2% |
| **History of chronic diseases** | | |
| Yes | 10.6% | 10.7% |
| No | 89.4% | 89.3% |
| **Family structure** | | |
| Living with both parents | 61.9% | 62.6% |
| Living with stepfamily | 16.5% | 16.6% |
| Living with single parent | 21.6% | 20.8% |
| **Parental alcohol consumption** | | |
| Low consumption | 76.2% | 76.5% |
| Moderate or high consumption | 23.8% | 23.5% |
| **Household smoking** | | |
| Yes | 45.1% | 44.8% |
| No | 54.9% | 55.2% |
| **Parental age category** | | |
| Teen parent | 13.1% | 13.2% |
| Age 20–34 | 81.8% | 81.7% |
| Age 35 or older | 5.1% | 5.0% |
| **Parental race** | | |
| White | 72.0% | 72.4% |
| Black | 14.2% | 14.2% |
| Hispanic | 9.9% | 9.5% |
| Other | 3.9% | 3.9% |
| **Parental stress** | | |
| Less stress | 96.8% | 96.7% |
| More stress | 3.2% | 3.3% |

| Characteristic | Eligible W5 biomarker sample (n = 4,640) | Final causal analytic sample (n = 4,050) |
|---|---|---|
| **Parental occupation** | | |
| Non-manual | 44.3% | 44.8% |
| Manual | 55.7% | 55.2% |
| **Parental education** | | |
| High | 65.6% | 67.0% |
| Low | 34.4% | 33.0% |
| **Migration background** | | |
| Non-immigrant | 87.8% | 88.7% |
| Immigrant | 12.2% | 11.3% |

#### J. Confirmatory Factor Analysis (CFA) and Scale Reliability

We conducted a CFA to assess the dimensional structure of the parenting responsiveness and demandingness constructs. The CFA was performed using the lavaan package in R version 4.5.1 using the weighted least squares mean and variance (WLSMV) adjusted estimator appropriate for ordered categorical items [6]. Two latent constructs were tested: responsiveness, reflecting parental warmth and support, and demandingness, reflecting parental control and monitoring. Internal consistency was subsequently assessed using Cronbach's alpha (α) and McDonald's omega (ω) using the psych package, also in R version 4.5.1.

For responsiveness, five items capturing perceived closeness, caring, warmth, communication quality, and overall relationship satisfaction loaded onto a single latent factor (standardized factor loadings 0.75 – 0.96). Model fit indices were as follows: Comparative Fit Index (CFI) = 0.995, Tucker-Lewis Index (TLI) + 0.991, Root Mean Square Error of Approximation (RMSEA) = 0.118, and Standardized Root Mean Square Residual (SRMR) = 0.066. The internal consistency for responsiveness was high ($\alpha = 0.85$, $\omega = 0.89$).

For demandingness, an initial CFA including seven items showed that the curfew item had weak loading on the demandingness factor. Given its comparatively poor psychometric performance, this item was excluded from the final measurement model. The retained six demandingness indicators, reflecting parental control over adolescents' daily decisions (friends, clothing, television hours, television programs, bedtime and food choices) loaded onto a single latent factor (standardized loadings 0.55 – 0.78). Model fit indicators were CFI = 0.975, TLI = 0.959, RMSEA = 0.051 and SRMR = 0.059. Reliability was acceptable ($\alpha = 0.63$, $\omega = 0.75$).

Consistent with recent methodological guidance, we did not evaluate fit against fixed cutoff values (e.g. CFI > 0.95, RMSEA < 0.06), which were developed for maximum-likelihood estimation with continuous data and are not valid for ordinal data analyzed with WLSMV[6]. Under WLSMV, CFI and TLI typically appear higher and RMSEA lower than under ML, so fit indices must be interpreted in context rather than relying solely on fixed cut-offs [6,7]. Given the strong and coherent loadings and good reliability, both constructs were considered adequately unidimensional and consistent with Baumrind's theoretical dimensions of responsiveness and demandingness. These constructs were therefore used as indicators to derive empirically defined parenting styles in the latent class analysis.

#### K. Latent Class Analysis (LCA)

We conducted a latent class analysis (LCA) to empirically identify patterns of parenting styles, based on Baumrind's two underlying constructs of parental responsiveness and demandingness. Responsiveness was measured using five adolescent-reported items (closeness, caring, warmth, communication and satisfaction with the relationship), and demandingness was measured using six items capturing parental rules or control (friends, clothing, television hours, television programs, bedtime and food). All items were treated as ordinal indicators.

Models specifying one to five latent classes were estimated in Stata 18 using the gsem package. Model fit was evaluated using the Akaike Information Criterion (AIC), Bayesian Information Criterion (BIC), adjusted BIC (aBIC) and entropy, while also considering the interpretability and stability of the resulting classes. Fit statistics for all candidate models are presented in Supplementary Table S4. The conditional probabilities of parenting styles for the two-class, three-class and five-class solutions are graphed in Supplementary Figures 3A – 3C.

The four-class solution provided the best balance of statistical fit and substantive interpretability. After selecting the optimal number of classes, we assigned each participant to the class with the highest posterior probability. These class assignments were used as the exposure in the causal analysis.

**Supplementary Table 4.** Model fit for 1 – 5 latent classes, n = 16,821. The best values that guided the choice of the model are in bold.

| Model | Log-likelihood | Resid. df | AIC | BIC | aBIC | Proportion of participants in smallest class (%) |
|---|---|---|---|---|---|---|
| 1 | -148566024 | 26 | 297318529 | 297313516 | 297316982 | N/A |
| 2 | -134637808 | 53 | 269827128 | 269813211 | 269819437 | 48.7 |
| 3 | -129081640 | 80 | 258163510 | 258164784 | 258160873 | 11.8 |
| **4** | **-126996488** | **107** | **254193045** | **254196351** | **254189964** | **11.0** |
| 5 | -126316338 | 134 | 252632817 | 252637123 | 252630281 | 5.1 |

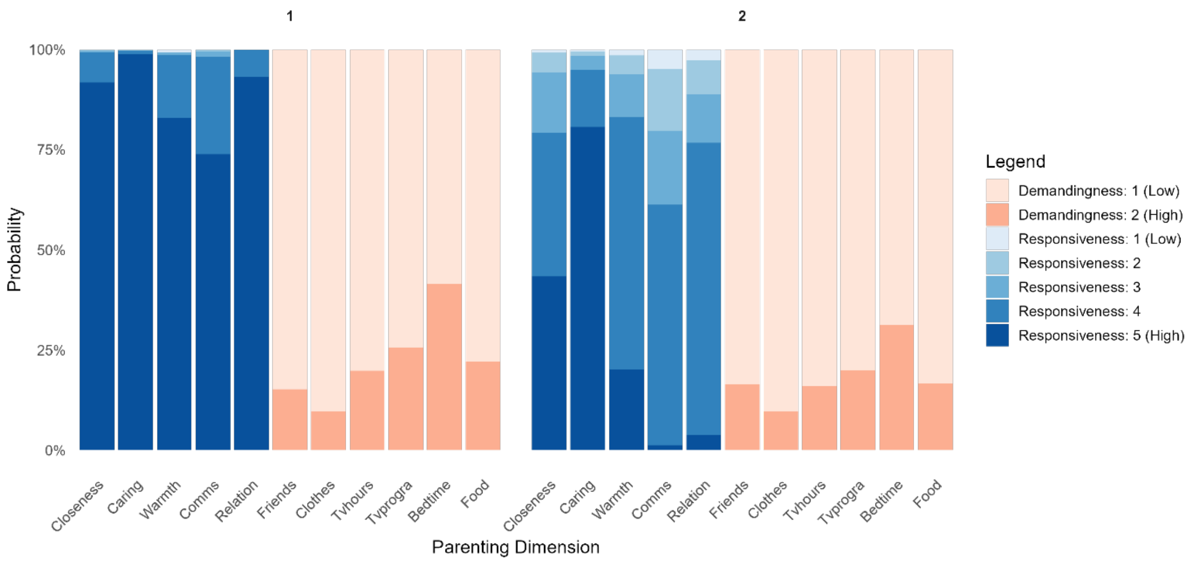

**Supplementary Figure 3A.** Two-class solution. Item-response probabilities by latent class for five responsiveness items (perceived closeness, caring, warmth, communication quality and relationship satisfaction) and six demandingness items (control over friends, clothing, television hours, television programs, bedtime and food).

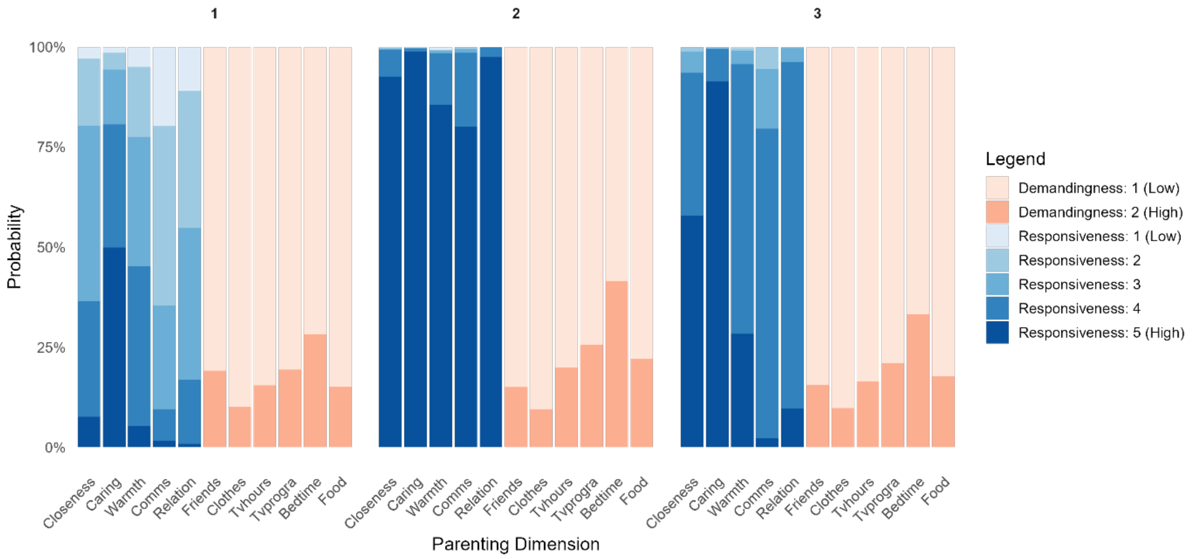

**Supplementary Figure 3B.** Three-class solution. Item-response probabilities by latent class for five responsiveness items (perceived closeness, caring, warmth, communication quality and relationship satisfaction) and six demandingness items (control over friends, clothing, television hours, television programs, bedtime and food).

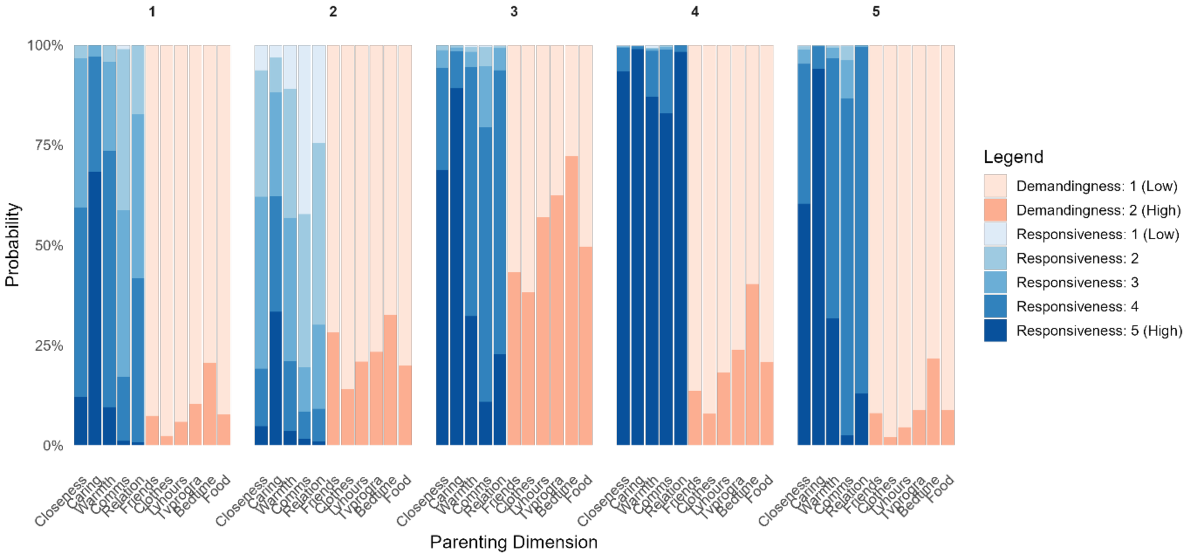

**Supplementary Figure 3C.** Five-class solution. Item-response probabilities by latent class for five responsiveness items (perceived closeness, caring, warmth, communication quality and relationship satisfaction) and six demandingness items (control over friends, clothing, television hours, television programs, bedtime and food).

#### L. Supplementary Analysis

We conducted a supplementary analysis where hypertension was the sole outcome.

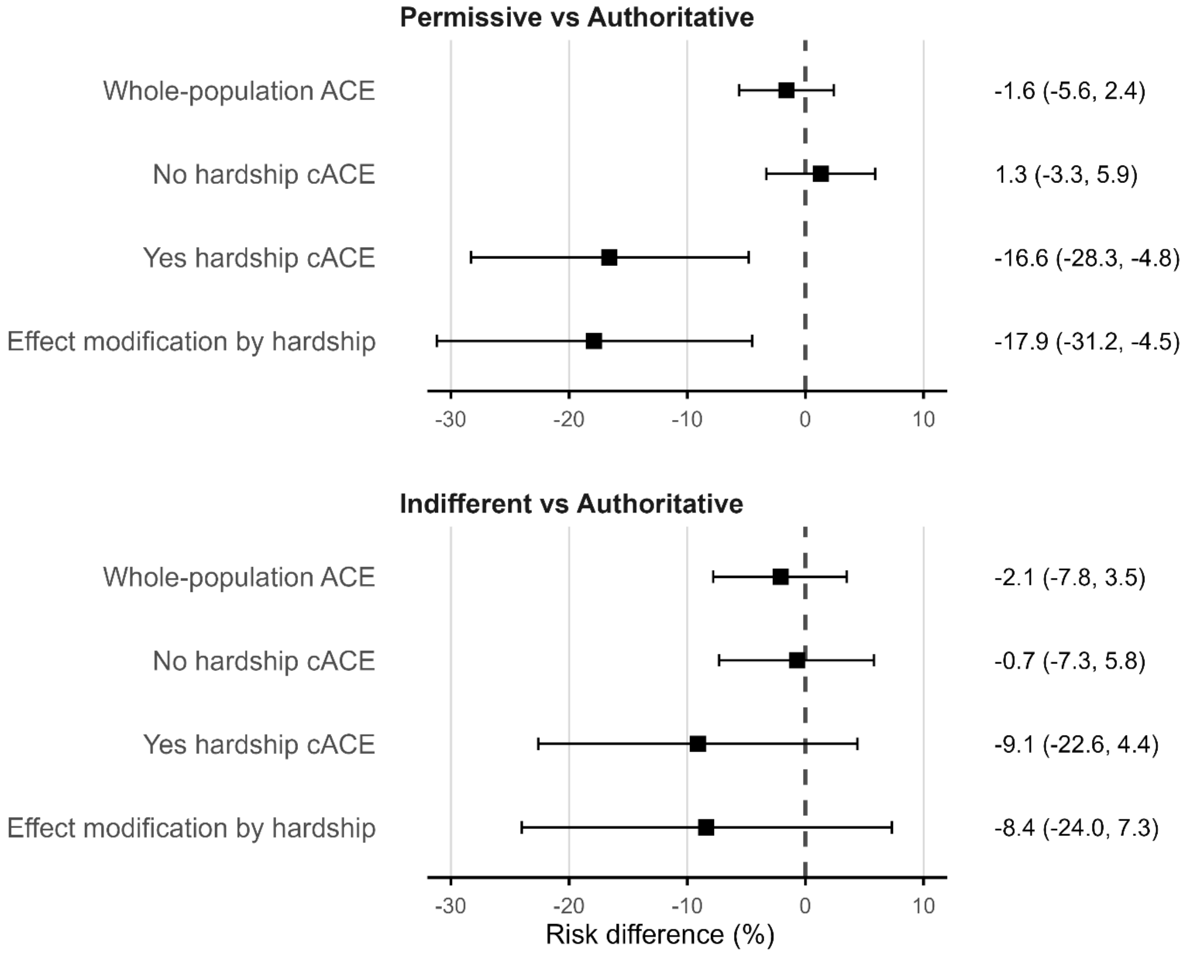

**Supplementary Figure 4.** Estimated whole-population average causal effect (ACE), conditional average causal effects (cACE, among adolescents without and with family financial hardship), and effect modification by hardship (difference between the two cACEs), for permissive vs. authoritative (top panel) and indifferent vs. authoritative parenting (bottom panel). Squares represent adjusted associational risk differences (%); their interpretation as causal effects relies on the assumptions stated in Methods. Error bars indicate 95% compatibility intervals. Point estimates and 95% compatibility intervals are also reported in the right-hand column.

#### M. Sensitivity Analyses

We conducted three sensitivity analyses to evaluate the robustness of our causal estimates.

##### a. Using Predefined Composite Parenting Styles

To assess whether our findings were dependent on the data-driven latent class structure, we redefined parenting style using a predefined composite approach based on Driscoll et al. (2007). This method dichotomizes parental responsiveness and demandingness and combines them into four theoretical categories: authoritative, authoritarian, permissive and indifferent. Using this predefined typology, we repeated the causal analysis, estimating risk differences stratified by family financial hardship and subsequently deriving effect modification estimates as the difference in conditional average causal effects (cACE) between adolescents with and without family financial hardship.

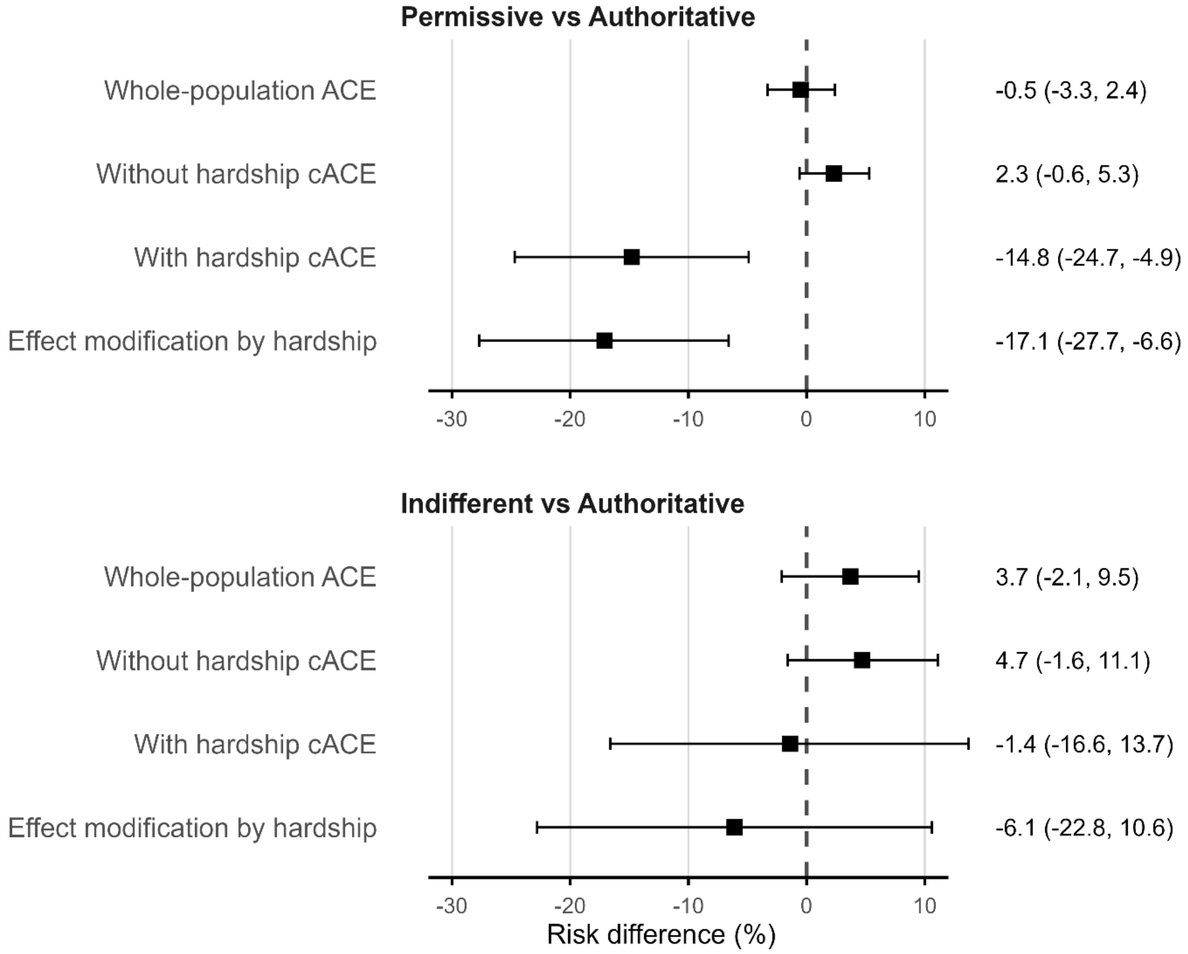

**Supplementary Figure 5.** Estimated whole-population average causal effect (ACE), conditional average causal effects (cACE, among adolescents without and with family financial hardship), and effect modification by hardship (difference between the two cACEs), for permissive vs. authoritative (top panel) and indifferent vs. authoritative parenting (bottom panel), using predefined composite parenting styles based on median splits of responsiveness and demandingness. Squares represent adjusted associational risk differences (%); their interpretation as causal effects relies on the assumptions stated in Methods. Error bars indicate 95% compatibility intervals. Point estimates and 95% compatibility intervals are also reported in the right-hand column.

#### b. Negative Control Exposure: Parental Seatbelt Use

To assess the potential impact of partially measured confounding, we conducted a negative control exposure analysis using parental seatbelt use while driving, reported in the Wave I parent survey. We selected seatbelt use because it is not expected to causally influence offspring cardiovascular or hypertension risk, but may capture underlying psychological or behavioural traits such as self-regulation, risk aversion or risk perception that could also relate to parenting style. These traits may not be fully measured by available covariates. Although our main analysis adjusted for parental psychosocial stress using self-reported general happiness as a proxy, this variable reflects only one aspect of such characteristics. Using parental seatbelt use in place of parenting style, we estimated the same risk difference contrasts to assess whether associations emerged that would be consistent with residual confounding.

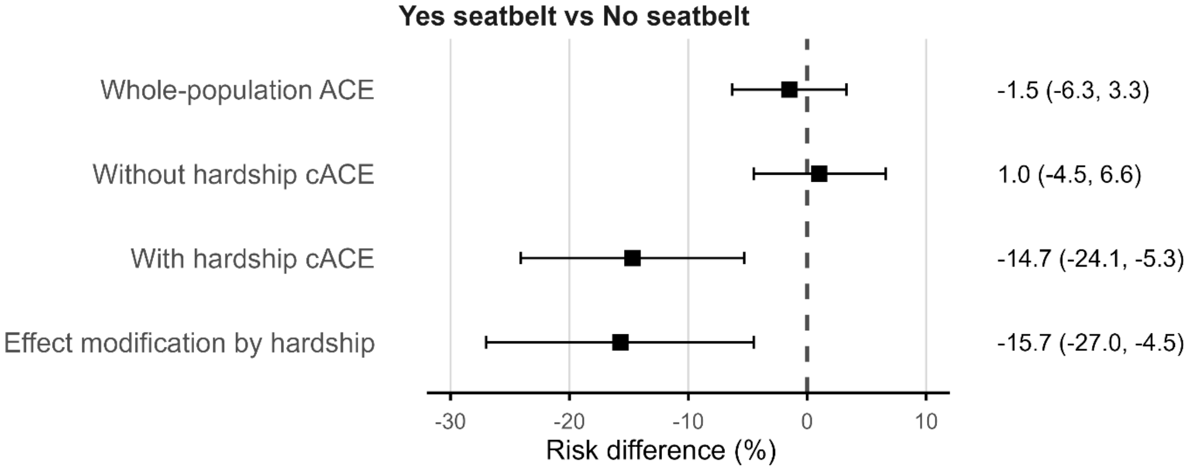

**Supplementary Figure 6.** Estimated whole-population average causal effect (ACE), conditional average causal effects (cACE, among adolescents without and with family financial hardship), and effect modification by hardship (difference between the two cACEs), for parental seat belt use vs. no use. Squares represent adjusted associational risk differences (%); their interpretation as causal effects relies on the assumptions stated in Methods. Error bars indicate 95% compatibility intervals. Point estimates and 95% compatibility intervals are also reported in the right-hand column.

#### c. Using Parental Education as an Alternative Effect Modifier

To examine whether results differ when using an alternative effect modifier, we repeated the analysis using parental education instead of family financial ability. Parental education was defined as the highest educational attainment of either parent, derived from the Wave I parent interview and adolescent survey, and categorized as high (at least one parent graduated from a college or university) versus low education. We repeated the causal analysis estimating risk differences stratified by parental education and subsequently derived effect modification estimates as the difference in cACEs between adolescents with low versus high parental education.

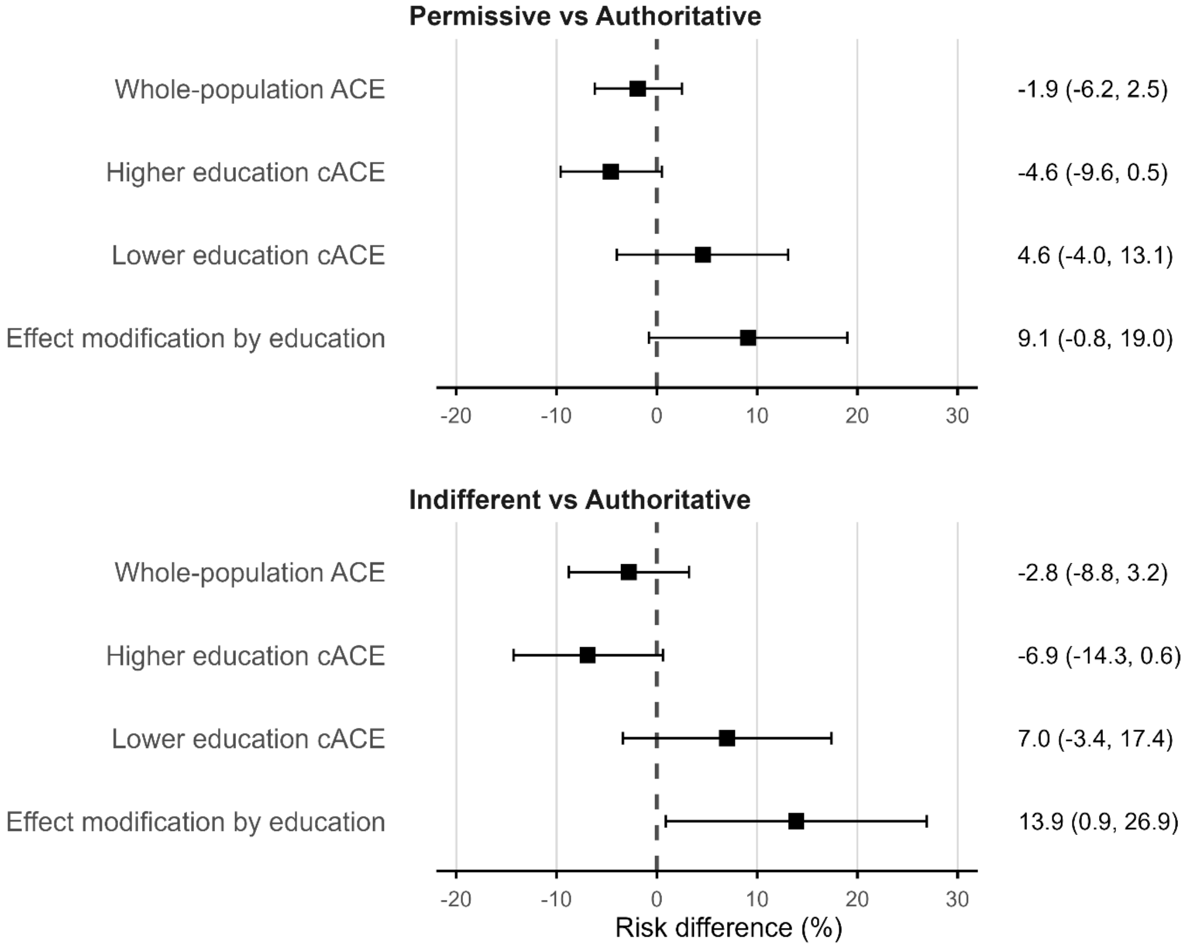

**Supplementary Figure 7.** Estimated whole-population average causal effect (ACE), conditional average causal effects (cACE, among adolescents with higher educated parents and with lower educated parents), and effect modification by parental education (difference between the two cACEs) for permissive vs. authoritative (top panel) and indifferent vs. authoritative parenting (bottom panel). Squares represent adjusted associational risk differences (%); their interpretation as causal effects relies on the assumptions stated in Methods. Error bars indicate 95% compatibility intervals. Point estimates and 95% compatibility intervals are also reported in the right-hand column.
